# Phylodynamics and migration data help describe HIV transmission dynamics in internally displaced people who inject drugs in Ukraine

**DOI:** 10.1101/2022.12.27.22283974

**Authors:** Ganna Kovalenko, Anna Yakovleva, Pavlo Smyrnov, Matthew Redlinger, Olga Tymets, Anna Korobchuk, Anna Kolodiazieva, Anna Podolina, Svitlana Cherniavska, Britt Skaathun, Laramie R. Smith, Steffanie A. Strathdee, Joel O. Wertheim, Samuel R. Friedman, Eric Bortz, Ian Goodfellow, Luke Meredith, Tetyana I. Vasylyeva

## Abstract

Internally-displaced persons are often excluded from HIV molecular epidemiology surveillance due to structural, behavioral, and social barriers in access to treatment. We test a novel field-based molecular epidemiology framework to study HIV transmission dynamics in a hard-to-reach and highly-stigmatized group, internally-displaced people who inject drugs (IDPWID). We inform the framework by Nanopore generated HIV *pol* sequences and IDPWID migration history. In June-September 2020, we recruited 164 IDPWID in Odesa, Ukraine, and obtained 34 HIV sequences from HIV-infected participants. We aligned them to publicly-available sequences (N=359) from Odesa and IDPWID regions of origin and identified 7 phylogenetic clusters with at least 1 IDPWID. Using times to the most recent common ancestors of the identified clusters and times of IDPWID relocation to Odesa, we infer potential post-displacement transmission window when infections likely to happen to be between 10 and 21 months, not exceeding 4 years. Phylogeographic analysis of the sequence data show that local people in Odesa disproportionally transmit HIV to the IDPWID community. Rapid transmissions post-displacement in the IDPWID community might be associated with slow progression along the HIV continuum of care: only 63% of IDPWID were aware of their status, 40% of those were in antiviral treatment, and 43% of those were virally suppressed. Such HIV molecular epidemiology investigations are feasible in transient and hard-to-reach communities and can help indicate best times for HIV preventive interventions. Our findings highlight the need to rapidly integrate Ukrainian IDPWID into prevention and treatment services following the dramatic escalation of the war in 2022.

**SIGNIFICANCE STATEMENT:** As human displacement is on the rise globally, it is crucial to develop ways in which infectious disease transmission can be monitored in displaced populations. We tested a new molecular epidemiology framework that relies on molecular epidemiology methods and portable HIV sequencing from samples collected from a hard-to-reach population of internally-displaced people who inject drugs (IDPWID). We show that by phylogenetically identifying potential HIV transmission clusters, estimating times of the clusters’ origin, and referencing these times against the time of IDPWID’s arrival to a new region, we can estimate an approximate window during an IDPWID’s displacement journey when HIV transmissions are likely to happen. Further analysis indicated that HIV is primarily transmitted from local populations to IDPWID.

## INTRODUCTION

An estimated 100 million people (over 1% of the global population) are currently forcibly displaced following economic, political, and climate instability (1). Internally displaced persons (IDPs, people who do not cross international borders) constitute the majority of forcibly displaced people. In 2021, 59.1 million IDPs were residing in low- and middle-income countries, 90% of whom have been displaced due to conflict and violence (2). In 2014, Ukraine, the country with the second-largest HIV epidemic in Europe (3), had between 1.4-2.1 million IDPs (Fig. 1A), the highest number in Europe, following the beginning of the war in eastern Ukraine (in Donetsk and Luhansk Oblasts) and the annexation of Crimea (Fig.1B) (4). Medical, economic, and social consequences of such large-scale displacement will have a long-lasting effect both on displaced and host populations.

**Figure 1.**
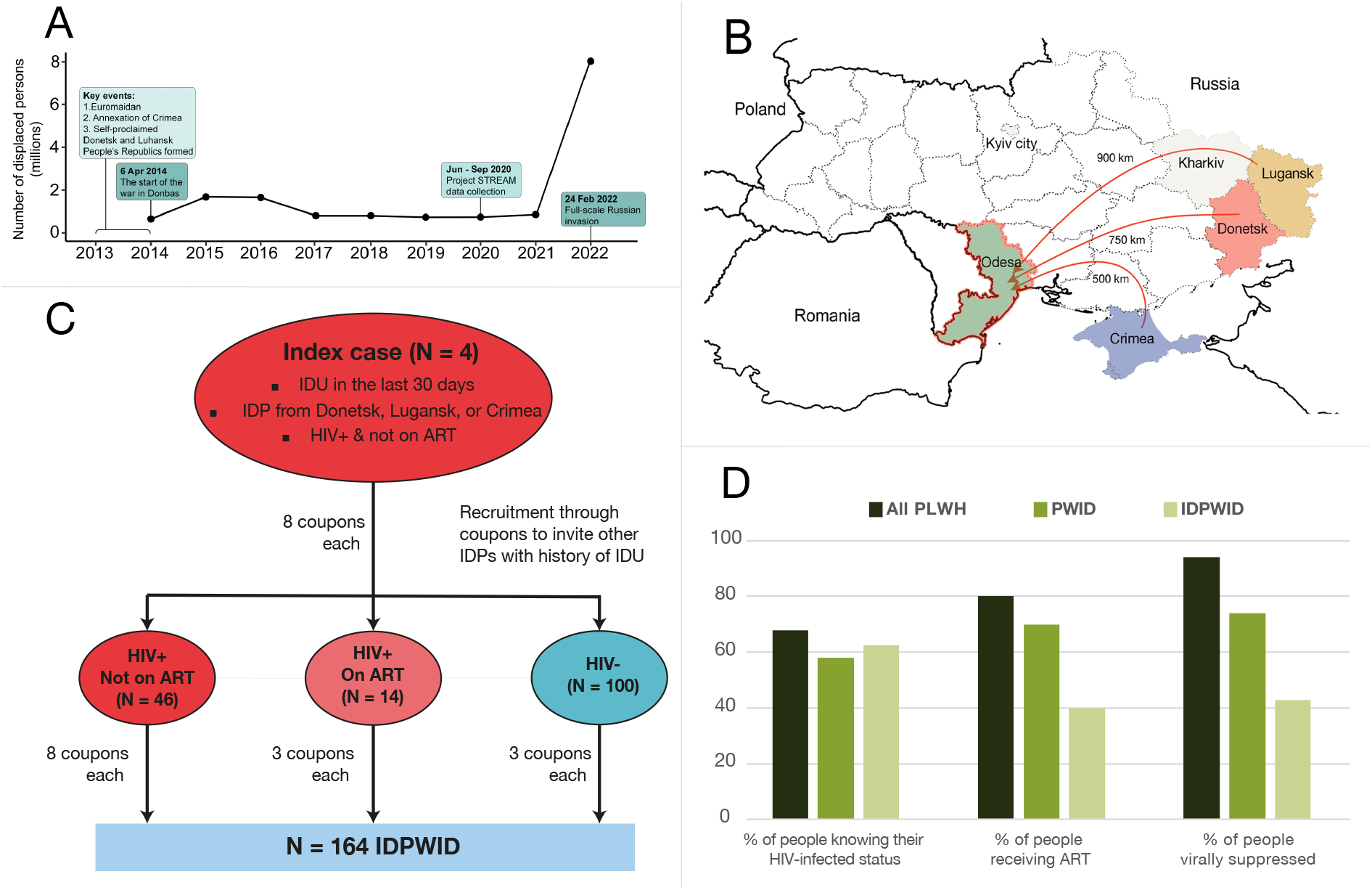
**A** – Timeline chart of key events of the Russia war on Ukraine and the number of internally displaced people in Ukraine over time; **B** – Map of Ukraine illustrating migration journeys. Beige color indicates regions that were often a place of settlement prior to arrival to Odesa; **C** - Recruitment flow diagram. IDU – injection drug use, ART – anti-retroviral treatment, IDPs – internally displaced persons, IDPWID – internally displaced people who inject drugs. **D -** Graphics of proportion of people engaged in each of the main stages of HIV continuum of care among key populations in Ukraine. PLWH – people living with HIV. Estimates for PLWH and PWID were obtained from (10) and (9).

Between 2014 and 2020, Odesa, a city on the Black Sea speculated to be the “birthplace” of Ukrainian HIV epidemic (5), has become home to nearly 40,000 Ukrainian IDPs (4). The prevalence of HIV and injection drug use (IDU) in IDPs resettled in Odesa remains unknown. Due to Russia’s stigmatizing approach to drug use (e.g., banning of medications for opioid use disorder), internally displaced people who inject drugs (IDPWID) disproportionately fled the occupied regions (6). Several successful HIV prevention interventions for PWID have been implemented in Odesa since 2013 (7, 8), but it is unknown whether IDPWID have had appropriate access to these preventive services upon relocation and whether HIV-infected IDPWID are enrolled in care. Generally, PWID in Ukraine progress less along the HIV care continuum compared to the general population: the latest available data for PWID in Ukraine showed that in 2017 58% of HIV-infected PWID were aware of their status, of whom 70% were on ART, and of whom 74% were virally suppressed (9), which was substantially lower than the same indicators for the general HIV-infected population in Ukraine in 2019 (68%-80%-94%, respectively) (10), though such comparisons are complicated by the absence of more recent data from PWID.

A number of factors mediate the relationship between displacement and HIV. For example, unstable housing post-displacement can be a barrier to accessing HIV care, and increasing the HIV vulnerability of displaced people (11). This might be exacerbated by behavioral risk factors such as IDU (12). IDPs’ social networks can play an important role: post-displacement social support networks can reduce HIV vulnerability in IDPs (13), but being in networks where IDU is prevalent puts displaced people at risk of injection initiation post-displacement (14).

Our previous phylogeographic analysis of HIV sequences available through routine drug-resistance testing illustrated that population displacement in Ukraine redistributed existing HIV infections and predisposed local outbreaks, particularly in PWID populations (15). While this analysis helped to reconstruct HIV transmission pathways and reveal key contextual insights, it likely undersampled IDPs, and particularly IDPWID, given that they are less likely to be in care and be selected for drug-resistance testing. This issue is not specific to Ukraine; global efforts to reinforce HIV molecular surveillance in the context of forced displacement are hindered by the scarcity of available sequencing data, primarily due to practical laboratory limitations concerning hard-to-reach mobile populations, such as IDPs. In the settings of large-scale forced displacement access to well-equipped laboratories is often limited. While field-based sequencing using lab-in-a-suitcase approach has been applied to other pathogens (16-18), it has not been utilized for HIV.

Here, we present results of the first study to combine rapid field-deployable Nanopore HIV sequencing conducted in a field-simulated environment and migration data collected from a hard-to-reach transient population (IDPWID). We integrate genetic and socio-behavioral information in a socio-molecular framework to describe HIV transmission and care dynamics among IDPWID following displacement in Ukraine. We suggest that by combining phylodynamic analysis with data about displacement histories, we can identify times when HIV transmissions are likely to happen in IDPWID. We also use phylogeographic analysis to infer directionality of HIV transmission between displaced and local populations.

## RESULTS

### Study population

A total of 164 IDPWID were recruited in a Spatial and Temporal Rapid Epidemiological Analysis in Migrants (STREAM) project in Odesa, Ukraine, in June-September 2020 using respondent-driven sampling (RDS) (19). We started recruitment with 8 seeds who were HIV-infected and not receiving ART IDPWID (details in Methods and Supplementary, Fig 1C). Most participants received 3 coupons to recruit peers; HIV-positive participants who were not receiving ART at the time of recruitment received up to 8 coupons. Most participants identified as male (81.1%) and arrived in Odesa between 2014-2015 (53.7%) (Table 1). The median participant age was 37 years old (range 20 – 63) at the time of enrolment. The median length of IDU experience was 10 years (range 2 – 40).

**Table 1.**
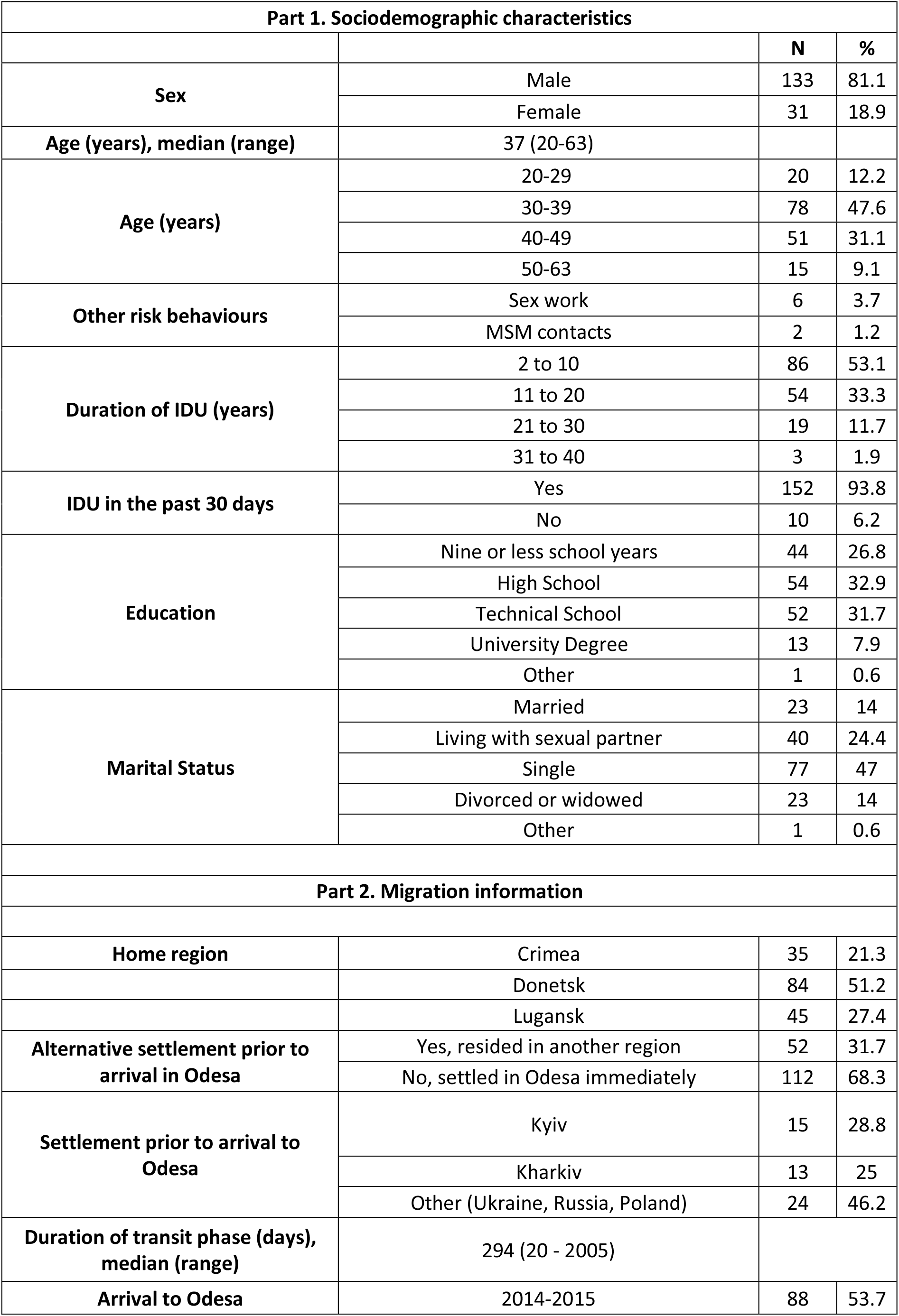
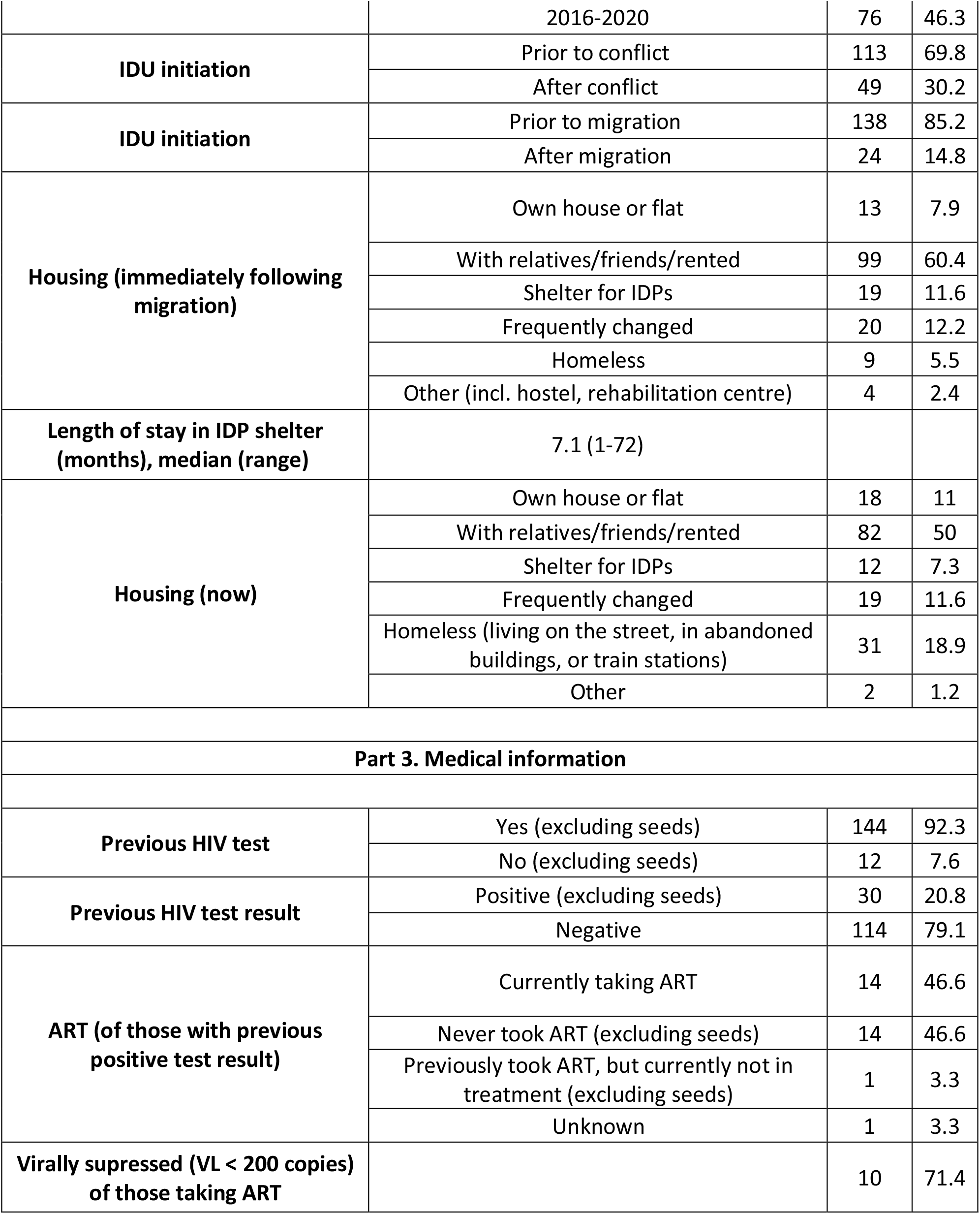
Characteristics of IDPWID recruited in Odesa in 2020

| **Part 1. Sociodemographic characteristics** |  |  |  |
| --- | --- | --- | --- |
|  |  | **N** | **%** |
| **Sex** | Male | 133 | 81.1 |
|  | Female | 31 | 18.9 |
| **Age (years), median (range)** | 37 (20-63) |  |  |
| **Age (years)** | 20-29 | 20 | 12.2 |
|  | 30-39 | 78 | 47.6 |
|  | 40-49 | 51 | 31.1 |
|  | 50-63 | 15 | 9.1 |
| **Other risk behaviours** | Sex work | 6 | 3.7 |
|  | MSM contacts | 2 | 1.2 |
| **Duration of IDU (years)** | 2 to 10 | 86 | 53.1 |
|  | 11 to 20 | 54 | 33.3 |
|  | 21 to 30 | 19 | 11.7 |
|  | 31 to 40 | 3 | 1.9 |
| **IDU in the past 30 days** | Yes | 152 | 93.8 |
|  | No | 10 | 6.2 |
| **Education** | Nine or less school years | 44 | 26.8 |
|  | High School | 54 | 32.9 |
|  | Technical School | 52 | 31.7 |
|  | University Degree | 13 | 7.9 |
|  | Other | 1 | 0.6 |
| **Marital Status** | Married | 23 | 14 |
|  | Living with sexual partner | 40 | 24.4 |
|  | Single | 77 | 47 |
|  | Divorced or widowed | 23 | 14 |
|  | Other | 1 | 0.6 |
| **Part 2. Migration information** |  |  |  |
| **Home region** | Crimea | 35 | 21.3 |
|  | Donetsk | 84 | 51.2 |
|  | Lugansk | 45 | 27.4 |
| **Alternative settlement prior to arrival in Odesa** | Yes, resided in another region | 52 | 31.7 |
|  | No, settled in Odesa immediately | 112 | 68.3 |
| **Settlement prior to arrival to Odesa** | Kyiv | 15 | 28.8 |
|  | Kharkiv | 13 | 25 |
|  | Other (Ukraine, Russia, Poland) | 24 | 46.2 |
| **Duration of transit phase (days), median (range)** | 294 (20 - 2005) |  |  |
| **Arrival to Odesa** | 2014-2015 | 88 | 53.7 |
|  | 2016-2020 | 76 | 46.3 |
| **IDU initiation** | Prior to conflict | 113 | 69.8 |
|  | After conflict | 49 | 30.2 |
| **IDU initiation** | Prior to migration | 138 | 85.2 |
|  | After migration | 24 | 14.8 |
| **Housing (immediately following migration)** | Own house or flat | 13 | 7.9 |
|  | With relatives/friends/rented | 99 | 60.4 |
|  | Shelter for IDPs | 19 | 11.6 |
|  | Frequently changed | 20 | 12.2 |
|  | Homeless | 9 | 5.5 |
|  | Other (incl. hostel, rehabilitation centre) | 4 | 2.4 |
| **Length of stay in IDP shelter (months), median (range)** | 7.1 (1-72) |  |  |
| **Housing (now)** | Own house or flat | 18 | 11 |
|  | With relatives/friends/rented | 82 | 50 |
|  | Shelter for IDPs | 12 | 7.3 |
|  | Frequently changed | 19 | 11.6 |
|  | Homeless (living on the street, in abandoned buildings, or train stations) | 31 | 18.9 |
|  | Other | 2 | 1.2 |
| **Part 3. Medical information** |  |  |  |
| **Previous HIV test** | Yes (excluding seeds) | 144 | 92.3 |
|  | No (excluding seeds) | 12 | 7.6 |
| **Previous HIV test result** | Positive (excluding seeds) | 30 | 20.8 |
|  | Negative | 114 | 79.1 |
| **ART (of those with previous positive test result)** | Currently taking ART | 14 | 46.6 |
|  | Never took ART (excluding seeds) | 14 | 46.6 |
|  | Previously took ART, but currently not in treatment (excluding seeds) | 1 | 3.3 |
|  | Unknown | 1 | 3.3 |
| **Virally suppressed (VL < 200 copies) of those taking ART** |  | 10 | 71.4 |

### HIV prevalence, testing history, and cascade of care

Previous HIV testing was reported by 91.5% (N = 150) of participants, of whom 24% (N=36) reported a previous positive test result. We removed the seeds (N=8) from further calculations because they were selected based on their HIV-status and engagement in care. Rapid testing as part of this study identified a total seroprevalence of HIV of 35.8% (N = 56 out of the 156 non-seed participants). Of those who tested positive for HIV by rapid test, 63% (N = 35) knew their HIV-infected status. Of those, 40.0% (N = 14) were on ART treatment at the time of the survey; 43% (N = 6) were virally suppressed (latest viral load measurement <50 copies/ml and no HIV amplification in this study) (Figure 1D).

### Migration journey, housing, and IDU

Most IDPWID had resided in Donetsk Oblast before 2014 (51.2%) and left it within a year from the beginning of the war in April 2014 (69.5%) (Figure 1A and 1B, Table 1). The majority of them immediately settled in Odesa following displacement (68.3%); of the rest, the most common regions for intermediate settlement prior to arrival in Odesa included Kyiv city (28.8%) and Kharkiv (25%) amongst other locations across Ukraine, Russia, and Poland. Those who immediately settled in Odesa reported that transit time between leaving their region of origin and arriving to Odesa was ≤2 days, whilst those settled elsewhere prior to arrival in Odesa had a median transit phase of 294 days (range 20 days – 5.5 years). The majority of IDPWID in our sample settled with relatives, friends, or in rented accommodation, both immediately following displacement and at the time of the study in 2020 (50% and 60.4%, respectively). Of those who resided in an IDP shelter immediately following displacement (11.6%), the median length of stay was seven months (range one month - six years). Over three times more participants reported homelessness in Odesa at the time of the study compared to immediately following displacement (18.9% vs. 5.5% of the sample, respectively). In this study, 30.2% (N=49) of IDPWID did not report IDU prior to the conflict, of whom 48.9% (N=24) reported starting IDU after relocation to Odesa. PWID in unstable housing situation at the time of the survey were more likely to report IDU in the last 30 days (*χ*^*2*^= 3.9, *p*=0·048).

### HIV sequence data

As a proof-of-principle, we conducted all wet laboratory work in a field-simulated environment at the Odesa Regional Virology Laboratory, utilizing a “lab-in-a-suitcase” approach (ARTIC Network) and using only portable and field-applicable equipment for Oxford Nanopore Technologies (ONT) sequencing as described elsewhere (16-18) (see Supplementary Text for more details). Of the 64 serum samples collected from participants who tested HIV-positive with a rapid HIV test, HIV was amplified in 47 samples which were further selected for library preparation and sequencing. No amplification in the other 17 samples as visualized by agarose gel electrophoresis was likely due to the low initial amount of the viral template RNA; 9 of those samples were from participants who were on ART. Of the 47 sequenced samples, 34 resulted in the assembly of partial *pol* gene sequences of protease (PR) and reverse transcriptase (RT) regions (1.1kb); the remaining 13 sequences had lower coverage depth (<20x threshold).

All 34 assembled sequences were assigned as HIV-1 subtype A1 (Supplementary Table 1). Only one major non-nucleoside reverse transcriptase inhibitor (NNRTI) DRM (K103N) was identified in 2 STREAM samples which constituted 6% or the total number of sequences (N = 34) and 7% of the treatment-naïve individuals for whom we obtained sequences (N = 28); 2 other NNRTIs (V179I and V179E) were identified in 12 other STREAM samples (35%) (see Supplementary Text for more details).

Additionally, we downloaded 359 reference sequences available through the Los Alamos National Laboratory (LANL) database sampled in Odesa, Donetsk, Luhansk, and Crimea (N = 252, 81, 16, and 10, respectively). Together with the 34 sequences generated in this study, the reference sequences formed a full dataset (N = 393) further used for phylogenetic analysis.

### Description of HIV transmission clusters

We inferred a Maximum Likelihood (ML) phylogenetic tree based on the sequences in the full dataset and identified 19 potential transmission clusters: 12 of them included reference sequences only and were described before (7), the other 7 included at least one IDPWID sequence from the STREAM project. The 7 STREAM clusters comprised 15 sequences (13 STREAM and 2 reference sequences): 6 dyads and 1 triad (Table 2 and Figure 2). Of the 13 STREAM participants whose sequences were in transmission clusters, 10 (77%) reported previous HIV testing post-displacement to Odesa, of those 9 (90%) knew about their HIV-positive status prior to participation in the STREAM study, and of those 6 had never taken ART (67%). Of the 3 participants who were on ART, two had their most recent viral load measurements at >200 copies/uL (49,300 and 14,500).

**Table 2.** Cluster composition and time to the most recent common ancestor (TMRCA) for HIV phylogenetic clusters

| Seq GenBank ID | Cluster | N of sequences | TMRCA | TMRCA 95% HPD | Time of IDPWID arrival to Odesa | Home region |
| --- | --- | --- | --- | --- | --- | --- |
| [OP899421, MT348863] | 1 | 2 | 2006 | 2000-2011 | May 2018 | Crimea |
| [OP899419, OP899437] | 2 | 2 | 2009 | Nov 2001-Jan 2016 | Aug 2014,<br>Jun 2015 | Donetsk,<br>Crimea |
| [OP899435, OP899443] | 3 | 2 | Jul 2017 | Feb 2013-Feb 2020 | Nov 2015,<br>May 2015 | Donetsk,<br>Lugansk |
| [OP899440, OP899441] | 4 | 2 | Sep 2013 | Nov 2007-Jan 2018 | Dec 2015,<br>Sep 2017 | Crimea,<br>Crimea |
| [OP899424, OP899410,<br>OP899423] | 5 | 3 | Nov 2017 | Oct 2014-Jan 2020 | Jan 2017,<br>Mar 2016,<br>Jan 2017 | Donetsk,<br>Donetsk,<br>Donetsk |
| [OP899431, MT348826] | 6 | 2 | 1997 | 1987-2007 | Mar 2020 | Donetsk |
| [OP899432, OP899434] | 7 | 2 | 2003 | 1992-2011 | Jul 2016,<br>Jul 2015 | Crimea,<br>Crimea |

**Figure 2.**
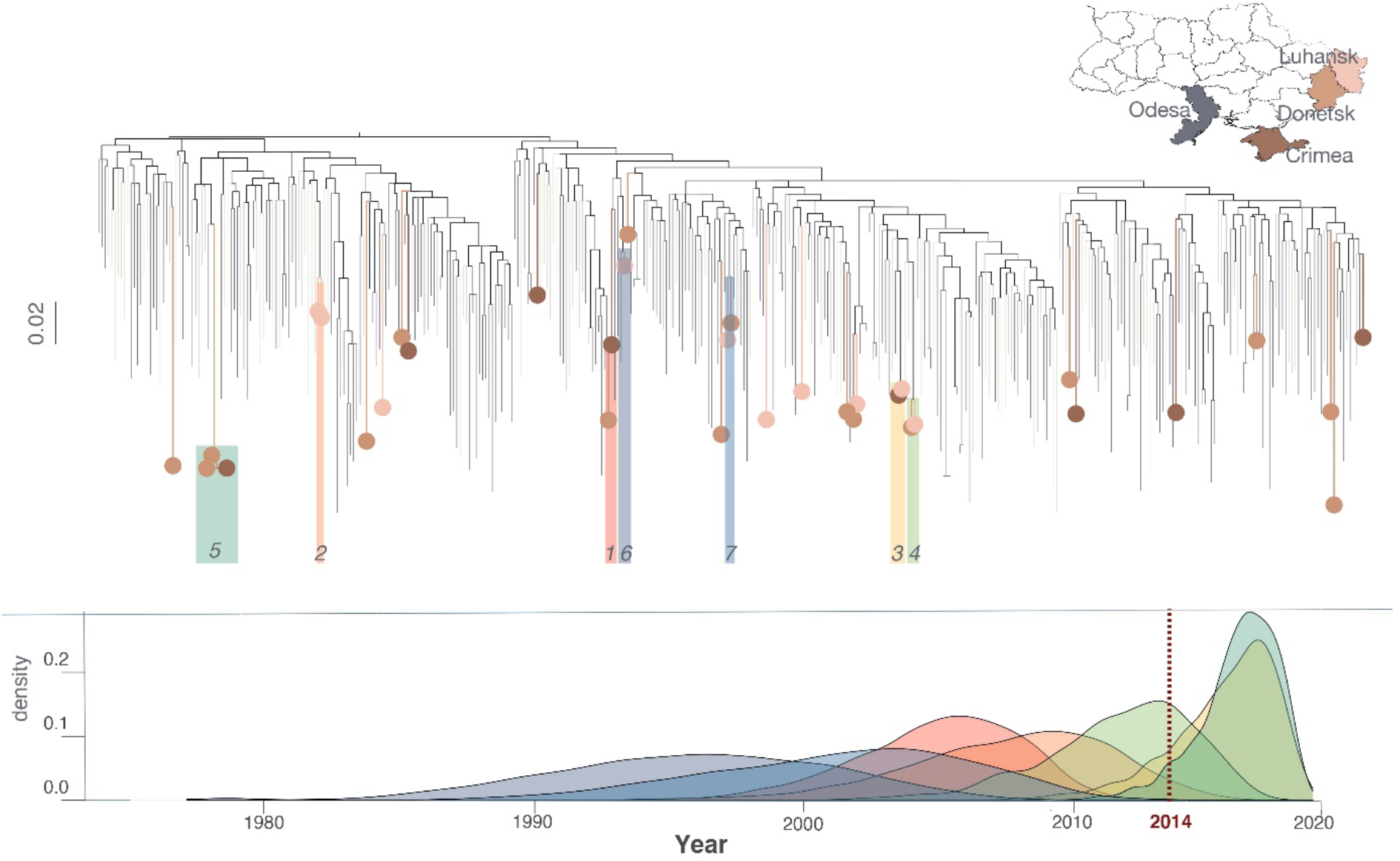
Maximum Likelihood phylogenetic tree based on the sequences in the full dataset. The colored circles on the tips correspond to the home regions of IDPWID sequenced in this study. Phylogenetic clusters are numbered and highlighted on the tree and TMRCA estimates for each cluster are presented on the plot below with corresponded colors (each color corresponds to a cluster as marked on the phylogenetic tree above). Scale bars indicate substitutions/site/year.

Clusters 3 and 5 likely originated post-conflict and after the time of the IDPWID arrival to Odesa. The time to the most recent common ancestor (TMRCA) for clusters 3 and 5 was inferred to be in July 2017 [95% Highest Posterior Density (HPD) February 2013 – February 2020] and November 2017 [October 2014 – January 2020], respectively. For cluster 3, the likely transmission window was between November 2015 and July 2017 (no later than February 2020) based on the time of arrival to Odesa of the last participant in the cluster and the cluster TMRCA. Participants in cluster 3 both had the K103N mutation (mutation sites were masked in all phylogenetic analyses). For cluster 5, the transmission likely happened between January 2017 and November 2017, based on the same criteria. The other five clusters are unlikely to represent direct transmission based on the fact that some participants come from different regions and cluster TMRCAs predate their arrival to Odesa; the TMRCAs for clusters 1, 2, 4, 6, and 7 were estimated to be in 2006 (2000–2011), 2009 (2001–2016), 2013 (2007–2018), 1997 (1987–2007) and 2003 (1992–2011), respectively.

### Phylogeographic analysis

We performed phylogeographic analysis to describe patterns of viral lineage movement between the IDPWID and local populations. The initial analysis included 252 reference Odesa sequences (“Local-Odesa” group which included sequences sampled from the general population and various transmission risk groups; at least 20% of these sequences were from PWID (7)) and 34 STREAM sequences (“IDPWID” group). This analysis showed that the absolute majority (99.7%, 95% HPD: 91%-100%) of viral lineage migration events (instances when a viral lineage moved from one group to another) were from the “Local-Odesa” to the “IDPWID” group. Since the number of sequences was substantially greater in the “Local-Odesa” group, we ran multiple sensitivity analyses to balance the number of sequences in both groups and match the number of sequences in the “Local-Odesa” and “IDPWID” groups (N=34 each and N=68 total for each subsample). For this purpose, analyses were repeated with the following subsampling approaches for the “Local-Odesa” group: 1) ten analyses with 34 random sequences from Odesa; 2) ten analyses only with Odesa sequences from PWID; 3) one analysis with 34 of the most recent sequences from Odesa (the most recent sequences from the “Local-Odesa” group included sequences collected in 2017, 2018, and 2019). In all these analyses, the “Local-Odesa” group remained the main viral lineage exporter – the average percent of viral lineage migration events across all 21 subsamples was 99% (95% BCI: 97-100%) (Supplementary Table 2).

In addition, to estimate the viral lineage movement pattern independently from the participants’ displacement status, we performed a permutation test. To that effect, in each of the ten subsamples from the first subsampling approach above we randomly rearranged (shuffled) displacement status assignments for the sequences. In this analysis, six out of the ten subsamples accounted on average for 99% (BCI: 95-100%) of viral lineage migration events from the “Local-Odesa” to the “IDPWID” group. Four out of 10 subsamples showed 99% (BCI: 94-100%) average proportion of viral lineage movements from the “IDPWID” to the “Local-Odesa” group.

## DISCUSSION

Here, we tested a framework for rapid epidemiological investigations in a hard-to-reach population often omitted from molecular epidemiological surveillance efforts and showed that a combination of phylodynamic analysis and information on migration history allows estimating the window when transmissions likely occurred post-displacement. IDPWID are particularly at risk for HIV soon after displacement and they are receiving HIV disproportionally from local communities.

Our phylogeographic analysis indicated consistent viral lineage movement from local populations to IDPWID consistent with the pattern observed in other migrants (20). In our analysis very few of the identified clusters included both sequences from IDPWID and local populations; either attributable to the low IDPWID sample size in our analysis or to limited mixing between the local population and IDPWID (21). IDPs often rely on support networks from their home towns for socialization and to address daily needs such as finding housing, creating tight networks post-displacement (22). Limited adaptation, recently reported for IDPs in Ukraine (23), could have created pockets of IDPWID within the Odesa PWID community that were not linked to HIV prevention. Such pockets would be susceptible to infections transmitted from the local population, which IDPWID need to rely on to procure drugs or injection equipment. Importantly, the directionality of HIV transmission from local to displaced population inferred through our phylogeographic analyses can contribute to dismantling the “anti-migration” narratives that picture migrants as a burden and a public health threat.

Our framework based on combining TMRCA estimates of phylogenetic clusters and migration histories of people in the clusters allowed us to estimate the window period during which HIV transmission occurred post-displacement to be between 10 and 21 months, not exceeding 4 years. This method complements other phylogenetically-based methods to estimate post-displacement HIV acquisition (24) by specifying the possible window for effective interventions. Using larger datasets or full-length genomes for analysis might allow further narrowing down of this window, which can inform the timeliness of prevention measures.

To enable rapid applications of this approach in environments without access to well-equipped laboratories, we performed all laboratory work in a field-simulated environment using only field-applicable equipment (a “lab-in-a-suitcase” approach) and ONT sequencing protocols. The equipment and workflow of a “lab-in-a-suitcase” have already proved to be robust and portable even with minimal power requirements, enabling effective implementation of viral genomic sequencing in resource limited settings (16-18). In war-affected settings and other settings with large-scale forced displacement, local laboratories might not be equipped for sequencing and molecular epidemiology research; in such settings “lab-in-a-suitcase” is feasible, reliable, and affordable approach. Even in situations when no laboratory facilities are available, the ‘lab-in-a-suitcase’ approach allows utilising other infrastructure or rely on mobile laboratory units. Moreover, as security conditions in such locations can deteriorate rapidly, also affecting further population migration, a “lab-in-a-suitcase” can be transported, assembled and disassembled easily and quickly, allowing the lab to be relocated immediately.

Although ONT sequencing initially reported high single-base error rate (1-4%) making this technology inapplicable in clinical HIV settings, improvements in the sequencing biochemistry and computational approaches for read accuracy, now render Nanopore protocols comparable to those of Sanger sequencing (25-28). While investigations in the use of MinION technology for identification of point-mutations and potential clinical applications are on-going, the current technology is likely sufficient for phylogenetic reconstruction, particularly that based on short sequences such as HIV PR-RT. Importantly, using ONT to produce HIV sequences in the field can help significantly reduce the previous lag between obtaining samples, genetic sequencing, and analysis of sequence data (16), and can provide near real-time evidence to inform HIV prevention services in rapidly developing situations, like the on-going war in Ukraine. Furthermore, it can make HIV sequencing more available and easier scalable in resource-limited environments.

Progression along the continuum of care, which affects post-displacement HIV transmission, was substantially lower in IDPWID (63% aware of their status/ 40% in treatment/ 43% virally suppressed) compared to that previously reported for non-displaced PWID in Ukraine (58%/70%/74%, respectively) (9) who were already behind the general population in Ukraine (68%/80%/94%, respectively) (10) (Figure 1D). Less than half of IDPWID were linked to care, indicating a huge gap in care engagement for Ukrainian IDPs, as we previously suggested [17]. Though normal RDS is recognized as an effective sampling approach for HIV studies in displaced populations (33), we recognize that ART uptake in our sample might be under-estimated given enhanced coupon distribution for HIV-positive participants who were not in treatment. However, the lower level of viral suppression in this cohort (less than half of those in treatment were virally suppressed) is an unbiased indicator of lower levels of HIV treatment engagement amongst IDPWID. The observed increase in homelessness, an independent risk factor for HIV acquisition for PWID [8], at the time of the study (18.9%), compared to the period immediately following displacement (5.5%), could also affect progression along the HIV care continuum. IDPs are likely to reside in high-risk environments with limited prevention and treatment resources, thus long-term housing solutions for IDPs could help minimize infection risks [6].

Many IDPWID in our sample initiated IDU after the beginning of the conflict (30.2%), half of those (48%) started injecting after they reached Odesa. Substance use amongst forcibly displaced people as a coping mechanism following conflict, violence, and migration stress is well documented in current literature [30] and will require a special focus in Ukraine, particularly in regions that are currently occupied by Russia where drug use is criminalized (34). While these data are subject to reporting biases, we expect these to be low in our sample, as is often the case for those for whom the recall events were more emotional or marked a transition point in their life, like war or forced displacement events for IDPWID [32].

As the war in Ukraine escalated to a full-scale invasion in February 2022 when Russia invaded multiple regions of Ukraine, resulting in the largest refugee crisis in Europe since World War II (Fig. 1A) (35), challenges in curbing the HIV epidemic in the country have reached unprecedented levels. ART drug supply disruptions, struggles in delivering prevention services, and huge reduction in HIV testing will have detrimental effect on the Ukrainian HIV epidemic (36, 37). For IDPWID, criminalization of drug use on the occupied territories and the customary exclusion of people who use drugs from HIV treatment services will create additional challenges (34). We show that integration of detailed sociodemographic and migration data alongside phylodynamic inference can help resolve transmission dynamics in this hard-to-reach populations and identify gaps in treatment and prevention efforts. Specifically, deciphering the infection timing in this study provides evidence for the necessity to scale up HIV prevention interventions within the timeframe of risk - the first 2 years following migration. Increased monitoring and understanding of the dynamics of infectious disease transmission networks, timing of transmission events, and risk factors unique to the experiences faced by IDPs is a crucial step in the development of effective health interventions.

## Data Availability

Nucleotide sequence data generated in this study are available in the GenBank database under the accession numbers: OP899410-OP899443.

## Acknowledgements

We acknowledge the assistance of public health workers from the non-governmental organisations “Alliance for Public Health” and “Way Home” in collection of epidemiological and behavioural data and biological samples, and their ongoing efforts of providing preventative measures and infection management to at risk populations in Ukraine. We also express our sincere gratitude towards study participants.

## Data availability

Nucleotide sequence data generated in this study are available in the GenBank database under the accession numbers: OP899410– OP899443.

## METHODS

### Study design and participants

This study was approved by the University of Oxford Tropical Research Ethical Committee (Reference: 530-20). Written informed consent was obtained from all participants. We used modified RDS technique (19) to recruit IDPWID starting with eight index cases, who were defined as people who 1) reported IDU in the last 30 days, 2) were internally displaced from Donetsk, Luhansk, or Crimea because of the war irrespective of their nationality, 3) were living with HIV, but were not receiving ART treatment at the time of study enrolment, and 4) were at least 18 years old. The number of distributed coupons differed depending on the HIV status of a participant (Figure 1C): the index cases and HIV-positive network members who were not in treatment at the time of the interview were given eight coupons to recruit other IDPs with history of IDU; HIV-positive IDPWID receiving ART and HIV-negative network members were offered three coupons to recruit other IDPWID. Recruitment was terminated after the field study period of three months was completed.

### Data Collection

Participants were interviewed by social workers from the non-governmental organization “Way Home” about their migration history, their sociodemographic and housing information, and screened for HIV using Wondfo® One Step HIV1/2 Whole Blood/Serum/Plasma. All participants who tested positive for HIV during the study were counselled by trained healthcare workers and linked to care. Whole blood samples (6 ml) were collected from all participants, and serum was isolated at the Odesa Regional Virology Laboratory and stored at -80°C.

### Statistical analysis

We defined current IDU status as IDU in the last 30 days (yes/no). We dichotomized the housing status into stable (living in one’s own house or flat, living with relatives or friends, and living in rented accommodation) and unstable (living in IDPs shelters, frequently changing accommodation, being homelessness and other housing types for example hostels and rehabilitation centers). We used a chi-square test to investigate whether there was a significant difference in recent IDU status by housing status post-displacement.

### Sequencing and bioinformatics

For library preparation viral RNA was extracted from 140 μL serum using QIAamp Viral RNA Minikit (Qiagen), following the manufacturer’s instructions. Extracted RNA was aliquoted and stored until use at -80°C. We adapted the primer set and a One-Step RT-PCR assay for genotyping all HIV-1 group M subtypes and circulating recombinant forms (CRF) developed by the Centers for Disease Control and Prevention (38, 39). The assay amplifies a fragment of 1,084 base pairs of the HIV-1 partial *gag-pol* polyprotein with protease and reverse transcriptase encoding regions (see Supplementary Text for details). The amplicons were purified with SPRI beads (x1 ratio), barcoded with native barcodes (EXP-NBD104 (1-12) and EXP-NBD114 (13-24)) and prepared with a ligation-based sequencing kit (SQK-LSK109) (ONT) according to the ARTIC Network nCoV-2019 Sequencing Protocol V3 LoCost (40). Final libraries were loaded onto new R9.4.1 flow cells (FLO-MIN106) and sequenced with a MinION Mk1B device. The field environment was simulated in the laboratory using a “lab-in-a-suitcase” approach developed by the ARTIC Network (18) where hydroponic grow tents were implemented to physically separate pre-PCR, post-PCR, and master mix work to prevent contamination of stock reagents or samples. Specifically, to simulate the “lab-in-a-suitcase” approach, only 12V batteries-powered library preparation equipment, such as vortexes, centrifuges, and miniPCR machines, was used. The MinION devices were powered from a USB3 port of a laptop. A curated equipment list is available on the ARTIC Network website (https://artic.network/ebov/ebov-seq-kit.html).

Reads were basecalled and demultiplexed with Guppy version 4.3.4. The reads were mapped to nucleotide positions 2100-3200 of HXB2 reference genome with minimap2. The reference was passed to Racon as a draft assembly and polished with the mapped reads. The resulting consensus was polished with Medaka. HIV subtype assignment was performed using REGA HIV-1 Subtyping Tool V3 (41).

### Phylogenetic analysis

We downloaded all publicly available through LANL sequences from Odesa and regions of IDPWID origin (Donetsk, Luhansk, and Crimea) with available sampling date information (Supplementary Text). 150 of the Odesa sequences on LANL came from the Transmission Reduction Intervention Study (TRIP) (7, 8, 42) and had associated information on IDU. We aligned all STREAM sequences to the reference data using Muscle algorithm (43) in Aliview (44), masked positions associated with drug-resistance (45), and used RAxML (46) to reconstruct Maximum Likelihood (ML) phylogenetic tree. Potential transmission clusters relevant to displacement were identified using ClusterPicker (47) and defined as clades with two or more sequences, at least one of which must be from IDPWID, that had within-cluster genetic distance <2.0%, which is within commonly-used thresholds for HIV clusters definition (48).

We inferred molecular clock phylogenetic trees from the full dataset and estimated the TMRCA of all identified transmission clusters using BEASTv1.10.8 (49) (BEAST analyses parameters are specified in the Supplementary Text). To quantify viral exchange between local Odesa population and IDPWID, we performed a discrete trait phylogeographic analysis using the tree distribution containing 2,000 trees previously estimated in BEAST. Sequences from Donetsk, Luhansk, and Crimea were pruned off the trees using PAUP as we were only interested in the dynamics between local and IDPWID populations in Odesa. We assigned STREAM sequences to the “IDPWID” group and all other sequences from Odesa were assigned to “Local-Odesa” group. To count the expected number of viral lineage migration events among transmission groups, we used a robust counting (Markov jumps) approach (15, 50). To control for the fact that we had a significantly higher number of sequences from Odesa, we ran three separate sensitivity analyses where we reduced the number of “Odesa” sequences to match the number of “IDPWID” sequences (details in the Supplementary Text).

## References

1. United Nations Higher Commissioner for Refugees: Ukraine, other conflicts push forcibly displaced total over 100 million for first time. 2022.

2. Internal Displacement Monitoring Centre, Global Report on Internal Displacement 2022. 2022.

3. European Centre for Disease Prevention and Control/WHO Regional Office for Europe, HIV/AIDS surveillance in Europe 2015. 2016, European Centre for Disease Prevention and Control: Stockholm.

4. United Nations Higher Commissioner for Refugees, Registration of Internal Displacement in Ukraine. 2021.

5. Saad MD, Shcherbinskaya AM, Nadai Y, Kruglov YV, Antonenko SV, Lyullchuk MG, et al. Molecular epidemiology of HIV Type 1 in Ukraine: birthplace of an epidemic. AIDS Res Hum Retroviruses. 2006;22(8):709–14.

6. Holt E. Conflict in Ukraine and a ticking bomb of HIV. Lancet HIV. 2018;5(6):e273–e4.

7. Vasylyeva TI, Zarebski A, Smyrnov P, Williams LD, Korobchuk A, Liulchuk M, et al. Phylodynamics Helps to Evaluate the Impact of an HIV Prevention Intervention. Viruses. 2020;12(4).

8. Smyrnov P, Williams LD, Korobchuk A, Sazonova Y, Nikolopoulos GK, Skaathun B, et al. Risk network approaches to locating undiagnosed HIV cases in Odessa, Ukraine. J Int AIDS Soc. 2018;21(1).

9. Sazonova Y, Kulchynska R, Sereda Y, Azarskova M, Novak Y, Saliuk T, et al. HIV treatment cascade among people who inject drugs in Ukraine. PLoS One. 2020;15(12):e0244572.

10. UNAIDS. Global AIDS Monitoring 2019: Ukraine 2019.

11. Stone J, Artenie A, Hickman M, Martin NK, Degenhardt L, Fraser H, et al. The contribution of unstable housing to HIV and hepatitis C virus transmission among people who inject drugs globally, regionally, and at country level: a modelling study. Lancet Public Health. 2022;7(2):e136–e45.

12. Arum C, Fraser H, Artenie AA, Bivegete S, Trickey A, Alary M, et al. Homelessness, unstable housing, and risk of HIV and hepatitis C virus acquisition among people who inject drugs: a systematic review and meta-analysis. Lancet Public Health. 2021;6(5):e309–e23.

13. Siriwardhana C, Ali SS, Roberts B, Stewart R. A systematic review of resilience and mental health outcomes of conflict-driven adult forced migrants. Confl Health. 2014;8:13.

14. Melo JS, Mittal ML, Horyniak D, Strathdee SA, Werb D. Injection Drug Use Trajectories among Migrant Populations: A Narrative Review. Subst Use Misuse. 2018;53(9):1558–70.

15. Vasylyeva TI, Liulchuk M, Friedman SR, Sazonova I, Faria NR, Katzourakis A, et al. Molecular epidemiology reveals the role of war in the spread of HIV in Ukraine. Proc Natl Acad Sci U S A. 2018;115(5):1051–6.

16. Faria NR, Quick J, Claro IM, Thézé J, de Jesus JG, Giovanetti M, et al. Establishment and cryptic transmission of Zika virus in Brazil and the Americas. Nature. 2017;546(7658):406–10.

17. Quick J, Loman NJ, Duraffour S, Simpson JT, Severi E, Cowley L, et al. Real-time, portable genome sequencing for Ebola surveillance. Nature. 2016;530(7589):228–32.

18. Brunker K, Jaswant G, Thumbi S, Lushasi K, Lugelo A, Czupryna A, et al. Rapid in-country sequencing of whole virus genomes to inform rabies elimination programmes [version 2; peer review: 3 approved]. Wellcome Open Research. 2020;5(3).

19. Heckathorn DD. Respondent-Driven Sampling: A New Approach to the Study of Hidden Populations. Social Problems. 1997;44(2):174–99.

20. Pimentel VF, Pingarilho M, Sole G, Alves D, Miranda M, Diogo I, et al. Differential patterns of postmigration HIV-1 infection acquisition among Portuguese immigrants of different geographical origins. AIDS. 2022;36(7).

21. Paraskevis D, Kostaki E, Nikolopoulos GK, Sypsa V, Psichogiou M, Del Amo J, et al. Molecular Tracing of the Geographical Origin of Human Immunodeficiency Virus Type 1 Infection and Patterns of Epidemic Spread Among Migrants Who Inject Drugs in Athens. Clin Infect Dis. 2017;65(12):2078–84.

22. Cantor D, Apollo, JO. Internal Displacement, Internal Migration and Refugee Flows: Connecting the Dots. London, UK: GCRF; 2020.

23. Singh NS, Bogdanov S, Doty B, Haroz E, Girnyk A, Chernobrovkina V, et al. Experiences of mental health and functioning among conflict-affected populations: A qualitative study with military veterans and displaced persons in Ukraine. Am J Orthopsychiatry. 2021;91(4):499–513.

24. Alvarez-Del Arco D, Fakoya I, Thomadakis C, Pantazis N, Touloumi G, Gennotte AF, et al. High levels of postmigration HIV acquisition within nine European countries. AIDS. 2017;31(14):1979–88.

25. Rang FJ, Kloosterman WP, de Ridder J. From squiggle to basepair: computational approaches for improving nanopore sequencing read accuracy. Genome Biol. 2018;19(1):90.

26. Zhang Y, Ma L. Application of high-throughput sequencing technology in HIV drug resistance detection. Biosafety and Health. 2021.

27. Gonzalez C, Gondola, J., Ortiz, AY, Castillo, J.M., Pascale, J.M., Martinez, A.A. Barcoding analysis of HIV drug resistance mutations using Oxford Nanopore MinION (ONT) sequencing. BioRXiv. 2018.

28. Sarkhouh H, Chehadeh W. CODEHOP-Mediated PCR Improves HIV-1 Genotyping and Detection of Variants by MinION Sequencing. Microbiol Spectr. 2021;9(2):e0143221.

29. Akinsete O, Hirigoyen D, Cartwright C, Schut R, Kantor R, Henry K. K103N mutation in antiretroviral therapy-naive African patients infected with HIV type 1. Clin Infect Dis. 2004;39(4):575–8.

30. Kirichenko A, Kireev D, Lopatukhin A, Murzakova A, Lapovok I, Saleeva D, et al. Prevalence of HIV-1 drug resistance in Eastern European and Central Asian countries. PLoS One. 2022;17(1):e0257731.

31. Palumbo PJ, Zhang Y, Fogel JM, Guo X, Clarke W, Breaud A, et al. HIV drug resistance in persons who inject drugs enrolled in an HIV prevention trial in Indonesia, Ukraine, and Vietnam: HPTN 074. PLoS One. 2019;14(10):e0223829.

32. Alliance for Public Health. Annual Report. Kyiv, Ukraine: ICF “Alliance for Public Health”; 2015.

33. Wirtz AL, Page KR, Stevenson M, Guillén JR, Ortíz J, López JJ, et al. HIV Surveillance and Research for Migrant Populations: Protocol Integrating Respondent-Driven Sampling, Case Finding, and Medicolegal Services for Venezuelans Living in Colombia. JMIR Res Protoc. 2022;11(3):e36026.

34. Roberts L. Surge of HIV, tuberculosis and COVID feared amid war in Ukraine. Nature. 2022;603(7902):557–8.

35. United Nations Higher Commissioner for Refugees. Refugees fleeing Ukraine (since 24 February 2022)*. 2022.

36. Holt E. Russia’s invasion of Ukraine threatens HIV response. Lancet HIV. 2022;9(4):e230.

37. Vasylyev M, Skrzat-Klapaczynska A, Bernardino JI, Sandulescu O, Gilles C, Libois A, et al. Unified European support framework to sustain the HIV cascade of care for people living with HIV including in displaced populations of war-struck Ukraine. Lancet HIV. 2022.

38. Inzaule S, Yang C, Kasembeli A, Nafisa L, Okonji J, Oyaro B, et al. Field evaluation of a broadly sensitive HIV-1 in-house genotyping assay for use with both plasma and dried blood spot specimens in a resource-limited country. J Clin Microbiol. 2013;51(2):529–39.

39. Zhou Z, Wagar N, DeVos JR, Rottinghaus E, Diallo K, Nguyen DB, et al. Optimization of a low cost and broadly sensitive genotyping assay for HIV-1 drug resistance surveillance and monitoring in resource-limited settings. PLoS One. 2011;6(11):e28184.

40. Quick J. nCoV-2019 sequencing protocol v3 (LoCost) V.3 Birmingham, UK 2020 [Available from: https://www.protocols.io/view/ncov-2019-sequencing-protocol-v3-locost-bp2l6n26rgqe/v3.

41. Pineda-Pena AC, Faria NR, Imbrechts S, Libin P, Abecasis AB, Deforche K, et al. Automated subtyping of HIV-1 genetic sequences for clinical and surveillance purposes: performance evaluation of the new REGA version 3 and seven other tools. Infect Genet Evol. 2013;19:337–48.

42. Nikolopoulos GK, Pavlitina E, Muth SQ, Schneider J, Psichogiou M, Williams LD, et al. A network intervention that locates and intervenes with recently HIV-infected persons: The Transmission Reduction Intervention Project (TRIP). Sci Rep. 2016;6:38100.

43. Edgar RC. MUSCLE: multiple sequence alignment with high accuracy and high throughput. Nucleic Acids Res. 2004;32(5):1792–7.

44. Larsson A. AliView: a fast and lightweight alignment viewer and editor for large datasets. Bioinformatics. 2014;30(22):3276–8.

45. Bennett DE, Camacho RJ, Otelea D, Kuritzkes DR, Fleury H, Kiuchi M, et al. Drug resistance mutations for surveillance of transmitted HIV-1 drug-resistance: 2009 update. PLoS One. 2009;4(3):e4724.

46. Stamatakis A. RAxML version 8: a tool for phylogenetic analysis and post-analysis of large phylogenies. Bioinformatics. 2014;30(9):1312–3.

47. Ragonnet-Cronin M, Hodcroft E, Hue S, Fearnhill E, Delpech V, Brown AJ, et al. Automated analysis of phylogenetic clusters. BMC Bioinformatics. 2013;14:317.

48. Hassan AS, Pybus OG, Sanders EJ, Albert J, Esbjornsson J. Defining HIV-1 transmission clusters based on sequence data. AIDS. 2017;31(9):1211–22.

49. Suchard MA, Lemey P, Baele G, Ayres DL, Drummond AJ, Rambaut A. Bayesian phylogenetic and phylodynamic data integration using BEAST 1.10. Virus Evol. 2018;4(1):vey016.

50. Minin VN, Suchard MA. Counting labeled transitions in continuous-time Markov models of evolution. J Math Biol. 2008;56(3):391–412.

