## Supplementary for "Phylodynamics and migration data help describe HIV transmission dynamics in internally displaced people who inject drugs in Ukraine"

### Sample data

All participants had complete data, apart from one participant for the variable "previous HCV test results" which is indicated in Table 1.

### Field-simulated environment using "lab-in-a-suitcase" approach

In order to make the workflow easily transferrable into a resource limited environment during possible refugee crises, we conducted all wet laboratory work in a field-simulated environment. The field-simulated environment involves the use of a "lab-in-a-suitcase" module (ARTIC network) which includes equipment with a small footprint, light weight, and minimal power consumption [16-18]. A "lab-on-a-suitcase" was deployed on the base of the Odesa Regional Virology Laboratory, the closest local lab to the study recruitment site. For this purpose, a separate space (room is divided into two functional zones by a plastic wall) in the building with the mains power supply and cold-chain equipment such as a fridge and freezer were provided. As at the time of the study, the lab was a part of the screening and surveillance COVID-19 testing program in the region, the space (room) was chosen to not interrupt the routine diagnostic work and was not included in the diagnostic unit. Three portable hydroponic tents were set up on standard desks to represent three separate hoods for the various steps of the library preparation: clean master mix tent (physically separated in a distant smaller zone 1), pre-PCR and post-PCR tents (zone 2). Additionally, one small hydroponic tent serving as a gel electrophoresis hood was set up in a separate room in the same building. A curated equipment list is available on the ARTIC network website (https://artic.network/ebov/ebov-seq-kit.html). A GELATO™ electrophoresis and visualization mini system (miniPCR bio; QP-1600-01) was used in this study but not included in the main equipment list on the website. Importantly, that before, after work, as well as between sample batches, cleaning/decontamination routine procedures should be performed in each tent (hood): all waste must be disposed of in sealed bags then the tent and all internal equipment must be cleaned (wiped) with 2 % sodium hypochlorite followed by 80% ethanol and then UV light cycle for at least 15 min to minimize the risk of nucleic acid contamination.

### RT-PCR

We used the primer set and a One-Step RT-PCR assay for genotyping all HIV-1 group M subtypes and circulating recombinant forms (CRF) developed by the Centers for Disease

Control and Prevention [38,39]. 20 µL of the extracted RNA was used with the SuperScript III One-Step RTPCR System with Platinum Taq High Fidelity DNA Polymerase (Invitrogen) in a 50 µL reaction and the following primer set: PRTM-F1, which is a mixture of primers F1a and F1b and is used as the forward primer, and RT-R1 reverse primer with concentration of 0.2 µM each of primers (PRTM-F1 and RT-R1) (Supplementary Table 3). The reaction mix was incubated with the following conditions: 50°C for 45 min and 94°C for 2 min as an initial RT cycle then 40 cycles of 94°C for 15 sec, 50°C for 20 sec, 72°C for 2 min and a final extension cycle at 72°C for 10 min using portable miniPCR machines. The RTPCR products – an approximately 1.1kb amplicons - were confirmed by 1% agarose gel electrophoresis using portable GELATO electrophoresis and visualization system.

**Mutations**

Fourteen (41,2 %) out of 34 participants were selected by NNRTI-RAM (2 - K103N; 10 - V179I; 2 - V179E). The K103N mutation is one of the most common NNRTI DRMs with a high-level resistance score and a very low or no virological response to ART [29]. While K103N mutation is reported to be common in Eastern European countries (found in 1.5-9.3% of samples depending on the country) [30], the prevalence in this study (2/34 or 5.9%) is higher than the 2.4% previously reported in another study of predominantly treatment-naïve PWID in Kyiv, Ukraine [31]; numbers in our study only rely on the observation of the mutation in two patients and are too low to allow comparisons in a statistical framework. Two (14,3 %) out of 14 participants who had DRMs reported to be ART-experienced (both had a V179I mutation): one was on ART at the time of sampling and another had taken ART in the past (Supplementary Table 1). All other participants (N=32) from whom HIV sequences were obtained, reported to be "treatment-naive" at the time of sampling; they have never undergone ART according to the participants' self-reports; these include the 12 participants whose HIV sequences had NNRTI-RAM.

Cluster analysis showed that of those 7 participants who were in HIV transmission clusters and reported no prior ART experience, two (in cluster 3) had an K103N NNRTI-RAM and another two (in cluster 4) had an V179E mutation. Among the three individuals in cluster 5, all had the V179I NNRTI-RAM but only one was on ART (latest viral load measurement 14500) at the time of recruitment. Another one STREAM participant with an V179I RAM was in cluster 1 together with a TRIP participant recruited in 2014. The higher prevalence of K103N, and the fact that participants with the K103N mutation were in a transmission cluster that originated post-displacement, might be evidence of the effect of the disruption of prevention and treatment services on HIV drug-resistance in IDPWID population, projected before [15, 32].

No DRM combinations were observed in this study.

**Sequence data and Phylogenetics**

We used reference sequences deposited from two sources: the Transmission Reduction Intervention Project (TRIP) conducted in 2013-2016 and from patients of the AIDS Centres in Odesa, Donetsk, Lugansk, and Crimea (host and origin regions for IDPWID in STREAM project) whose samples were sequenced to assess drug resistance mutations at the Gromashevskyi Institute of Epidemiology between 2012 and 2019. Our data set comprises HIV-1 subtype A pol genetic sequences corresponding to HXB2 nucleotide positions 2101–3200. Further details on the TRIP data collection and genetic sequencing procedures can be found in the original publication [7,42].

We used RAxML [46] to reconstruct Maximum Likelihood (ML) phylogenetic tree under General Time-Reversible nucleotide substitution model with gamma-distributed rate variation among sites and proportion of invariable sites (GTR + G + I) and ran a bootstrap analysis with 1000 replicates.

### BEAST analyses

To estimate the times to most recent common ancestor of the identified transmission clusters, we estimated molecular clock phylogenetic trees using BEAST1.10 [49]. We used Bayesian SkyGrid population dynamics model with a cut-off value of 1970 under the GTR+G+I nucleotide substitution model. For molecular clock inference, we used a lognormal relaxed molecular clock model with a continuous-time Markov chain (CTMC) prior on the mean (constrained to the values above 1 × 10−3) and an exponential prior on the standard deviation (mean = 0.333). We also used a lognormal distribution prior for the age of the root of the tree (mean = 1974, standard deviation = 5). Four Markov chain Monte Carlo (MCMC) chains were run for 300 × 106 iterations and upon convergence were combined using LogCombiner with 10% removed as burn-in for each chain. The estimated trees from the four chains were also combined and subsampled at a lower frequency resulting in a tree distribution of 2,000 trees.
